# Safety and tolerability of intravenous liposomal GM1 in patients with Parkinson disease: A single center open-label clinical phase I trial (NEON trial)

**DOI:** 10.1101/2024.09.05.24313105

**Authors:** Stefan Halbherr, Stefanie Lerch, Sebastian Bellwald, Petra Polakova, Bettina Bannert, Marie Roumet, Roch-Philippe Charles, Martin A. Walter, Corrado Bernasconi, Camille Peitsch, Pascal C. Baumgartner, Céline Kaufmann, Heinrich P. Mattle, Alain Kaelin-Lang, Andreas Hartmann, Michael Schuepbach

## Abstract

**Background:** Parkinson disease (PD) is a chronic progressive neurodegenerative disorder leading to motor and non-motor impairment often resulting in severe loss of quality of life. There are symptomatic treatments without effect on the progression of PD. A disease-modifying treatment that could ideally stop the neurodegenerative process is direly needed. Monosialotetrahexosylganglioside (GM1) is a promising molecule with neuroprotective effects in preclinical models of PD and has yielded encouraging results in patients with PD in a randomized placebo-controlled trial. Talineuren (TLN) is a liposomal formulation of GM1 that has been shown to cross the blood-brain barrier in animals. We assessed the safety and pharmacokinetics of TLN in patients with PD.

**Methods and Findings:** We prospectively enrolled 12 patients with PD into a single-center, open-label phase I trial to assess the safety and tolerability of weekly infusions with Talineuren. The maximum suitable dose of Talineuren was determined by dose escalation in three patients. Subsequently, these and nine further patients received weekly infusions at the maximum suitable dose of Talineuren over two months (1 patient stopped prematurely). All adverse events were continuously assessed as the primary objective and coded according to the Medical Dictionary for Regulatory Activities (MedDRA®). Clinical manifestations of PD were assessed as secondary outcomes using the Movement Disorders Society Unified Parkinson’s Disease Rating Scale (MDS-UPDRS) including a levodopa challenge test at baseline and end. In addition to weekly history taking, scales to measure mood, behaviour, quality of life, sleepiness, non-motor symptoms of PD, and cognition were used. Dose escalation identified a maximum suitable weekly dose of 720 mg. Overall, 304, mostly mild adverse events occurred. Twenty-three were considered related to the study treatment. Very mild to severe acute infusion reactions at the 2nd, 3rd, or 4th administration of TLN within the first minutes of the infusion occurred in seven patients. All reported back or neck pain. Other acute infusion reactions were urticaria, plethora, nausea, and chest pain. These adverse reactions disappeared within minutes of stopping the infusion and did not recur when Talineuren administration was resumed at a very low rate. Beyond the 4th administration, infusions could be given at increased rates up to 370 ml/h and no acute reaction occurred anymore. The mechanism of this acute infusion reaction remains unclear. Some patients reported mild dizziness for a few hours after Talineuren following many but not all administrations throughout the study. Non-motor symptoms of PD, motor parkinsonian signs off medication, and quality of life improved significantly during the treatment phase, including the MDS-UPDRS total score (mean decrease 11.09±10.47 points; p=0.006) and the PDQ-39 summary index (mean decrease 2.91±2.25 points, p =0.002). Dopaminergic medications remained stable during the study.

**Conclusion:** Talineuren is safe and well-tolerated in general. This prospective phase I trial revealed non-allergic habituating acute infusion reactions at the 2nd, 3rd, or 4th treatment that can be prevented by a slower rate of infusion. Importantly, the exploratory results suggest a consistent improvement of signs and symptoms of PD.

## Introduction

Parkinson disease (PD) is a chronic progressive neurodegenerative disorder with motor signs and non-motor symptoms. Its clinical hallmark is Parkinsonism, i.e., bradykinesia associated with rest tremor or rigidity (1). Although the motor signs remain the primary defining feature of PD, vegetative, behavioural, and cognitive symptoms may have a predominant effect on quality of life, and often precede motor signs (2). Histologically, PD is characterized by alpha-synuclein containing neuronal inclusions called Lewy bodies that propagate through the brain resulting in impaired function of numerous systems (3,4). Some manifestations of PD can symptomatically be relieved with medication or stereotactic procedures, but despite considerable ongoing research efforts (5), there is currently no known disease-modifying treatment available for PD. The most important therapeutic potential currently consists of medications for the substitution of dopamine loss due to degeneration of dopaminergic neurons in the substantia nigra. However, over decades of disease progression levodopa-resistant symptoms become the main cause for disability. A treatment to slow down or even halt the pathological process in PD is direly needed.

The glycosphingolipid GM1 (monosialotetrahexosylganglioside) is an important component of the cell membrane of neurons that is diminished in patients with PD (6). GM1 has shown neurotrophic and neuroprotective properties in preclinical research (7), and decreased levels of GM1 in patients with PD may contribute to the pathogenesis (8). Studies in animal models of PD have shown recovery with GM1 treatment (9), and a randomized double-blinded placebo-controlled clinical study in 77 patients treated with twice daily subcutaneous administration of 100 mg GM1 resulted in a significant benefit compared to placebo over 120 weeks (10) Administration and therapeutic effects of GM1 may be improved by using a neurotropic nanoparticle carrier. Talineuren (TLN) is a liposomal formulation consisting of the active pharmaceutical ingredient (API) GM1 at 6 mg/ml and the carrier liposomes consisting of sphingomyelin and cholesterol. We conducted a phase I safety trial in 12 PD patients receiving weekly TLN infusions over 8 weeks minimum.

## Methods

### Study Design

In this single-centre, open-label phase I interventional trial we enrolled 12 patients with PD to assess the safety, tolerability, and preliminary efficacy of liposomal GM1 as add-on medication with weekly intravenous infusions. Three patients received weekly ascending doses for 14 weeks to establish the highest well-tolerated dose (dose escalation group [DE]). The highest tested weekly dose of the API was 720 mg based on previous use of GM1 in humans (11) and was reached in all three patients. DE patients and 9 additional patients (dose consolidation group [DC]) received thereafter weekly TLN with an API dose of 720 mg for 8 weeks between December 13, 2021 and June 20, 2022. One of these patients stopped participation prematurely. One month after the last administration of TLN, a final safety follow-up visit was performed. Antiparkinsonian medication was kept stable if possible. In two patients with bilateral subthalamic nucleus stimulation, stimulation remained on and parameters unchanged throughout the study.

An independent data safety monitoring board (DSMB) evaluated safety data when the 3 DE patients had reached a GM1 dose of 180 mg and after completion of the dose escalation. The trial was approved by the local ethics committee (Kantonale Ethikkommission Bern) and the competent authority October 30, 2021. The trial is registered at the US National Institute for Health (ClinicalTrials.gov NCT04976127). The trial was conducted according to the principles of the Declaration of Helsinki and Good Clinical Practice guidelines. Talineuren is provided by the manufacturer and sponsor of the study, InnoMedica Switzerland AG. Talineuren is produced under GMP conditions at InnoMedica’s Nanofactory in Marly (Freiburg, Switzerland).

### Patients

Patients aged 40-80 years were eligible if they were diagnosed with PD according to British brain bank criteria (12), had a Hoehn and Yahr Stage 0 – 2.5 on medication (13) and stable PD treatment for at least 4 weeks. To exclude major cognitive deficits, a score >25 on the Montreal Cognitive Assessment (14) was required. Patients were carefully selected to exclude medical, psychological, and behavioural problems that may have interfered with their compliance for study participation. Patients provided written informed consent and fulfilled all eligibility criteria.

### Dose escalation (DE)

In patients 1-3 dose escalation was started at 6 mg GM1 and then increased weekly to 12 mg, 60 mg, followed by increases of 60 mg/week up to 720 mg. TLN was provided as a concentrated liposomal suspension containing cholesterol, sphingomyelin, and the active pharmaceutical ingredient GM1 in 30ml vials (180 mg GM1) in a phosphate buffered solution. Talineuren was diluted in phosphate buffered saline with a final concentration of 6 mg/ml GM1. TLN was added to 250 ml NaCl 0.9% and administered with 250 ml/h with a perfusor. The speed of infusion could be increased up to 370 ml/h if tolerated but reduced if necessary. Three days after the infusion each weekly dose increase of the TLN had to be cleared by an internal safety monitoring committee (MS, RPC, MW) by assessing clinical symptoms and lab values (Appendix 1). The second and third patient could start with the lowest dose with a delay of 1 week after confirmation of safety in the first patient. This 1+2 schedule was maintained for each weekly increase to allow for modification of the increasing of doses if needed according to a predefined dose modification matrix. All 3 patients had two clinical and lab assessments each week during the 14 weeks of the dose escalation phase. The average of the three maximal tolerated doses was defined as maximal suitable dose (MSD) for the dose consolidation (DC) part of the trial.

### Dose consolidation (DC)

After DE, the initial 3 and additional 9 patients received weekly infusions of TLN with an API dose of 720 mg and clinical and lab safety assessments over 8 weeks except for one patient who stopped participation early. A final safety assessment was performed one month after the last administration of TLN.

### Pharmacokinetics

Pharmacokinetic parameters were determined for GM1 from individual concentration time profiles obtained after the first intravenous infusion administration of 720 mg GM1 in the 9 DC patients. Blood samples were drawn before the infusion started and after the start of the infusion (250 ml/h, 370 ml total volume) at 5 minutes, 1 hour, 4, 8, 24, 48, 72, and 96 hours. Samples were immediately centrifuged and frozen at -70°C. GM1 concentration was measured using liquid chromatography with tandem mass spectrometry.

Noncompartmental pharmacokinetic analysis was performed using Phoenix WinNonlin version 8.0 [Pharsight Corporation, Mountain View, CA, USA] using the intravenous infusion dosing option. Pharmacokinetic variables were estimated from the plasma concentration versus time curves: tmax (time to reach the maximum plasma concentration read directly from the plasma concentration-time curve), Cmax (maximum plasma concentration read directly from the plasma concentration-time curve), AUC0-∞ (area under the plasma concentration-time curve from time point zero to infinity, estimated from AUC0-t + Ct/λz, where t is the last sampling time with a concentration above the limit of quantification and λz is the terminal elimination rate constant, estimated by log-linear least squares regression of the plasma concentration versus time data in the terminal phase; AUC0-∞ was calculated according to the linear trapezoidal with linear/log interpolation rule), t½: (apparent terminal half-life calculated as ln2/λz), CL: (apparent clearance calculated as dose divided by AUC0-∞), Vz (apparent volume of distribution calculated as dose divided by λz * AUC0-∞).

### Outcome measures

The primary outcome was safety defined as the occurrence of adverse events. Safety assessments included a full medical history at baseline, a full general physical medical and neurological examination at baseline, a final assessment and last follow-up, and detailed unstructured interviews at each visit and if needed by phone. A narrative description of adverse events and their duration, intensity, seriousness, and relation to TLN were recorded by the site investigators. Adverse events were coded by an independent expert with the Medical Dictionary for Regulatory Activities (MedDRA®).

Adverse events were considered unrelated to TLN if a plausible other explanation for the observation was available. Otherwise, adverse events without plausible causal relation to TLN but lacking a different explanation were considered to be unlikely related to TLN. Safety data were weekly reviewed by an internal safety board (MS, RPC, MAW) during the dose escalation period. Any out-of-range laboratory result was counted as an adverse event.

Parkinsonian motor signs and non-motor symptoms were further explored weekly using the Movement Disorders Society-Unified Parkinson’s Disease Rating Scale (MDS-UPDRS) (15) parts 1 (non-motor experiences of daily living), 2 (motor experiences of daily living), 3 (motor examination), and 4 (motor complications). The MDS-UPDRS-2 was assessed for best and worst condition in the preceding week in patients with motor fluctuations. The MDS-UPDRS-3 was assessed before and after each administration of TLN and also at baseline and final assessment in a levodopa challenge test (LCT). For the LCT patients paused their dopaminergic medications for at least 12 hours before the assessment “off” medication. The usual morning dose of levodopa equivalence plus 50 mg levodopa was then given as liquid formulation of levodopa/benserazide (in 1 patient levodopa/carbidopa) for the ensuing assessment “on” medication. Levodopa equivalent daily doses were noted weekly and calculated according to standard procedures (16). The number of PD-related non-motor symptoms was assessed with the Non-Motor Symptoms Questionnaire (NMSQuest) (17) at baseline and weekly during the course of the trial. Questionnaires at baseline and final assessment included the Parkinson’s disease Questionnaire-39 (PDQ-39) (18) for disease-related quality of life, the Epworth Sleepiness Scale (19), the Montreal Cognitive Assessment (MoCA (20) for mental performance, the Beck Depression Inventory (BDI) (21) to assess mood, and the Starkstein Apathy Scale (22).

### Statistical analysis

After completion of data entry, data validation and cleaning were performed. Data analysis was started. All patients enrolled in this study received at least one dose of study medication and were considered in the safety and tolerability analysis.

PK analysis plasma concentrations and PK parameters were analyzed for the 9 patients of the consolidation group and reported in a descriptive fashion. Descriptive statistics include arithmetic mean, SD, minimum, median, maximum, geometric mean, and coefficient of variation of arithmetic and geometric means.

Other study outcomes including scores measuring the Parkinsonian motor signs and non-motor symptoms, disease related quality of life, levodopa equivalent daily dose, and mental performance were assessed in the 11 patients who received at least two full-dose infusions and completed at least the MDS-UPDRS at these two visits. For patients included in the escalation part we used as baseline value the values assessed at the start of the trial (i.e. before the start of the escalation part).

We used summary statistics to describe the outcomes values at baseline, after 8 weeks of treatment, and the observed changes. For each outcome, the significance of the change from baseline was assessed using a paired Wilcoxon signed-rank test. No correction for multiplicity was applied.

Few outcomes including the blood lab values of cholesterol (total, LDL, HDL, ratio total/HDL), triglycerides, and apolipoprotein B were defined, post-hoc, as additional outcomes. For these outcomes, we used summary statistics to describe the weekly assessed values. At each time point, the significance of the change from baseline was assessed using a paired Wilcoxon signed-rank test.

## Results

### Trial population

There were no screening failures. Twelve patients (including three women) were recruited. At inclusion the median age was 65 (range 46-75) years old and had had motor parkinsonian signs for 7.9±5.2 (range 2-19, median 6.5) years. Three patients had only akinetic-rigid signs and nine patients also rest tremor. All patients had been clinically diagnosed with PD. For the patient characteristics see Table 1.

**Table 1.** Patient characteristics.

| Characteristic | Overall, N = 12 | Dose consolidation, N = 9 | Dose escalation, N = 3 |
| --- | --- | --- | --- |
| Age at registration (years) |  |  |  |
| Median (range) | 65.0 (46.0–75.0) | 65.0 (51.0–75.0) | 63.0 (46.0–68.0) |
| Sex - Female, n (%) | 3 (25%) | 2 (22%) | 1 (33%) |
| Height (cm) |  |  |  |
| Median (range) | 172.0 (159.0–183.0) | 172.0 (160.0–183.0) | 174.0 (159.0–182.0) |
| Weight (kg) |  |  |  |
| in | 78.5 (60.0–99.0) | 78.0 (64.0–99.0) | 88.0 (60.0–99.0) |
| BMI (kg/m<sup>2</sup>) |  |  |  |
| Median (range) | 26.2 (21.6–32.7) | 25.8 (21.6–30.8) | 26.6 (23.7–32.7) |
| Hoehn and Yahr stage at screening, n (%) |  |  |  |
| Stage 0 (no signs of disease) | 0 (0%) | 0 (0%) | 0 (0%) |
| Stage 1 (unilateral involvement only) | 3 (25%) | 2 (22%) | 1 (33%) |
| Stage 1.5 (unilateral and axial involvement) | 5 (42%) | 4 (44%) | 1 (33%) |
| Stage 2 (Bilateral involvement without impairment of balance) | 4 (33%) | 3 (33%) | 1 (33%) |
| Stage 2.5 (Mild bilateral disease with recovery on pull test) | 0 (0%) | 0 (0%) | 0 (0%) |
| Drugs |  |  |  |
| L-DOPA, n (%) | 12 (100%) | 9 (100%) | 3 (100%) |
| Non-ergot-derived dopamine receptor agonist, n (%) | 8 (67%) | 6 (67%) | 2 (67%) |
| MAO-B Inhibitor, n (%) | 4 (33%) | 3 (33%) | 1 (33%) |
| COMT inhibitor, n (%) | 3 (25%) | 3 (33%) | 0 (0%) |
| NMDA agonist, n (%) | 1 (8.3%) | 1 (11%) | 0 (0%) |
| Other, n (%) | 1 (8.3%) | 1 (11%) | 0 (0%) |
| LEDD (mg) |  |  |  |
| Median (range) | 650.0 (340.0–1,275.0) | 750.0 (340.0–1,275.0) | 550.0 (375.0–1,010.0) |

### Safety

### Dose escalation

Weekly dose escalation of TLN from 6 to 720 mg GM1 i.v. at a rate of 250 ml/h was well tolerated without major adverse effects. Mild neck and lumbar pain occurred for 15 minutes in one patient during the second (12 mg) and reappeared very mildly for a few minutes during the third (60 mg) infusion with TLN. Otherwise, dose escalation was unremarkable for all three patients and reached 720 mg with good tolerance. One patient reported transient beneficial effects of TLN treatment on sleep, mood, and general energy levels during dose escalation lasting longer with increasing doses. At the end of the dose escalation, the subjective beneficial effects of the weekly infusions lasted almost a week in this patient.

### Pharmacokinetics

After first intravenous application of TLN with an API dose of 720 mg, repeated blood sampling showed the plasma peak of TLN in the sample taken 4 hours after start of the infusion in all but one patient in whom the peak was reached at 1 hour after start of the infusion. This patient was the patient who had the severe acute infusion reaction at the second administration. Pharmacokinetic variables are given in Table 2.

**Table 2.** Pharmacokinetic variables for GM1 after i.v. infusion administration of 720 mg GM1 in the dose consolidation patients group (n = 9).

| n=9 | C<sub>BL</sub><br>(ng/ml) | C<sub>max</sub><br>(ug/ml) | t<sub>½</sub><br>(h) | AUC<sub>0-∞</sub><br>(h*ug/ml) | V<sub>z</sub><br>(ml) | CL<br>(ml/h) |
| --- | --- | --- | --- | --- | --- | --- |
| Mean | 52.8 | 212 | 15.0 | 5'011 | 4'188 | 210 |
| SD | 30.8 | 153 | 6.48 | 4'035 | 2'666 | 113 |
| Minimum | 0 | 84 | 10.2 | 1'920 | 2'020 | 49 |
| Median | 65.4 | 147 | 12.6 | 3'860 | 3'290 | 187 |
| Maximum | 84.5 | 582 | 30.7 | 14'700 | 10'000 | 375 |

C<sub>BL</sub>: baseline plasma concentration before TLN administration. C<sub>max</sub>: maximum plasma concentration read directly from the plasma concentration-time curve. t<sub>½</sub>: apparent terminal half-life. AUC<sub>0-∞</sub>: area under the plasma concentration-time curve from time point zero to infinity. V<sub>z</sub>: apparent volume of distribution. CL: apparent clearance.

### Dose consolidation

During the dose consolidation phase, acute infusion reactions occurred in 6 patients shortly after the start of the 2^nd^ (6 patients), 3^rd^ (1 patient), and 4^th^ (1 patient) administration of TLN (see below). One patient with a severe infusion reaction did not receive any further TLN treatment. During all following treatments with TLN, no acute infusion reactions occurred. The maximum dose of GM1 720 mg/week could be maintained throughout the study.

### Adverse events

Overall, 304, mostly mild adverse events occurred. No serious adverse event was reported in this study. Twenty-three adverse events were affirmatively related to the study treatment. The causal relation to the study treatment was considered probable in 17 (all mild), possible in 100, and unlikely in 123 observations. Forty-one adverse events were unrelated to the study treatment.

Adverse events definitively or probably related to TLN can be separated into a group of immediate mild to severe infusion reactions that abated after halting the administration of TLN (see below), and a group of more diffuse, always mild reactions often occurring with some hours’ delay after the TLN infusion. The latter group comprises mild tension headache (n=3), nausea (n=1), and arterial hypertension during emotional tension (n=1) during the infusion, and dizziness (n=6), inner tension and tremulousness (n=2), and tension headache (n=4) occurring within hours after the infusion, spontaneously remitting within less than a day. One patient suffered from mildly depressed mood during the withdrawal of TLN after the dose escalation phase, partly because he missed the beneficial effects of the weekly infusions. This symptom remitted spontaneously before TLN was resumed in the consolidation phase of the study.

Acute infusion reactions occurred in one patient during dose escalation (2^nd^ and 3^rd^ administration), and in six patients during dose consolidation, always within minutes of the 2^nd^ administration of TLN. In two patients, an acute infusion reaction was observed at the 3^rd^ administration, in one very mildly during the 4^th^ administration. Symptoms occurred after as little as 3 mg of liposomal GM1 was administered and as early as within 1 minute after start of the infusion. Symptoms built up at variable speeds and with mild to severe intensity among different patients. All symptoms abated within minutes after stopping the infusion. Infusion could then be resumed at a low speed (10 ml/h) without recurrence of symptoms. In most cases, infusion speed could be increased swiftly up to the planned 250 ml/h except in one patient who required several hours for the 2^nd^ infusion. However, later the patient tolerated administrations of TLN at 250 ml/h without any adverse events. In one patient, the acute infusion reaction at the beginning of the 2^nd^ administration quickly led to severe back, neck, and chest pain. According to the study protocol, the patient had to be removed from the study and could not be re-exposed at a lower infusion rate despite the patients’ explicit request to continue study participation. The acute infusion reactions (Table 3) included lower back, neck, and chest pain as the most common symptom; lower back pain was present in all patients with an acute infusion reaction. Lower back pain was very mild in two patients during three administrations and did not require the infusion to be halted. One patient had itching during 2^nd^, 3^rd^, and 4^th^ administration of TLN, and urticaria on the trunk and the thighs (Table 3) during the 3^rd^ infusion. Tryptase was normal before start of TLN and within 30 minutes of the appearance of urticaria. Clemastine 2 mg was given i.v. and TLN could be resumed without worsening of symptoms. Urticaria disappeared within a day and did not recur during the following administrations of TLN. After the last occurrence of an acute infusion reaction, patients received 19 (n=1, dose escalation), 6 (n=4), and 4 (n=1) infusions without experiencing an acute infusion reaction. One patient was withdrawn from the study (Figure 1). In the remaining 5 patients overall 68 TLN infusions were given without an acute infusion reaction.

**Fig 1:**
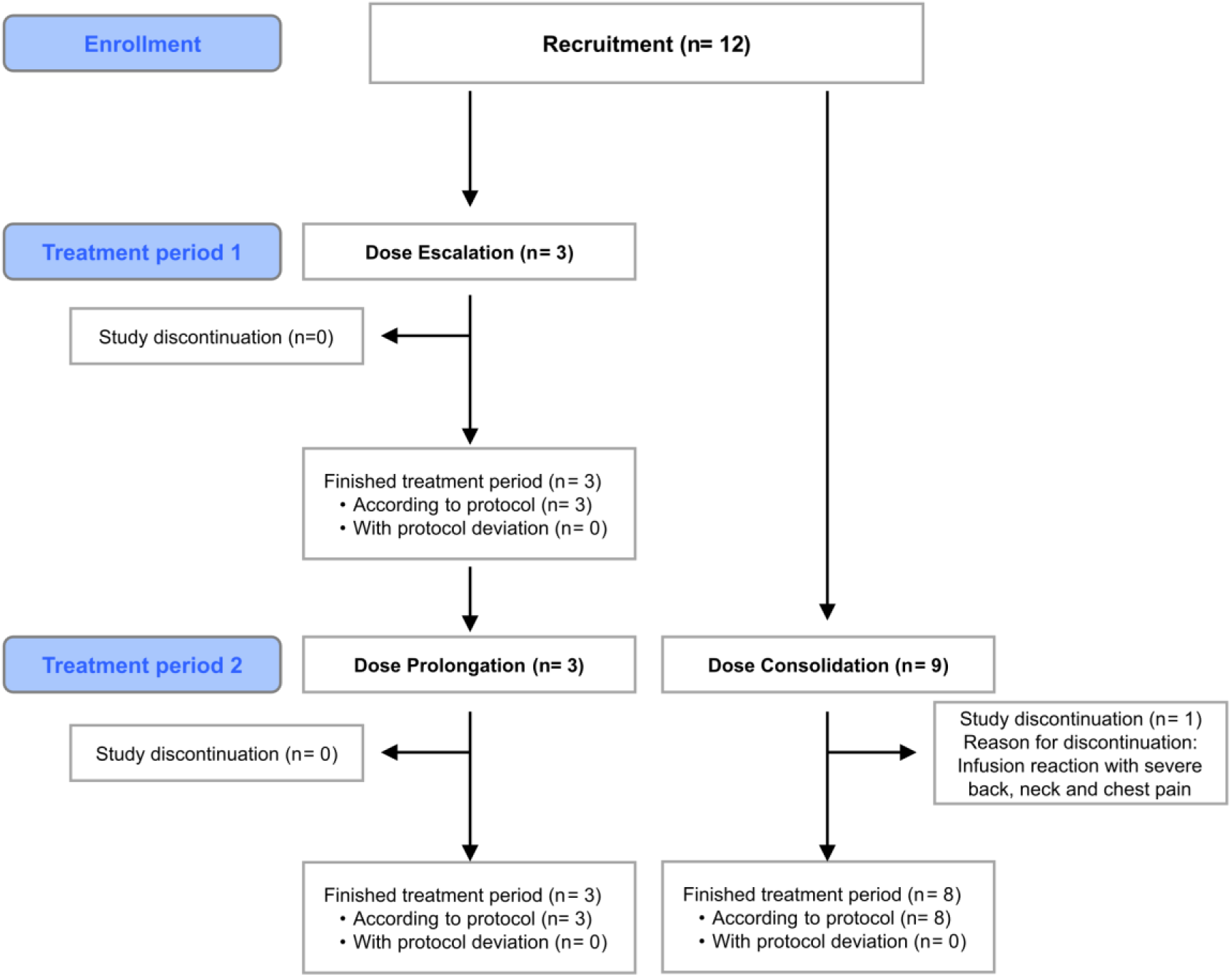
**CONSORT diagram of dose escalation group (with prolongation) and dose consolidation group.**

**Fig 2.**
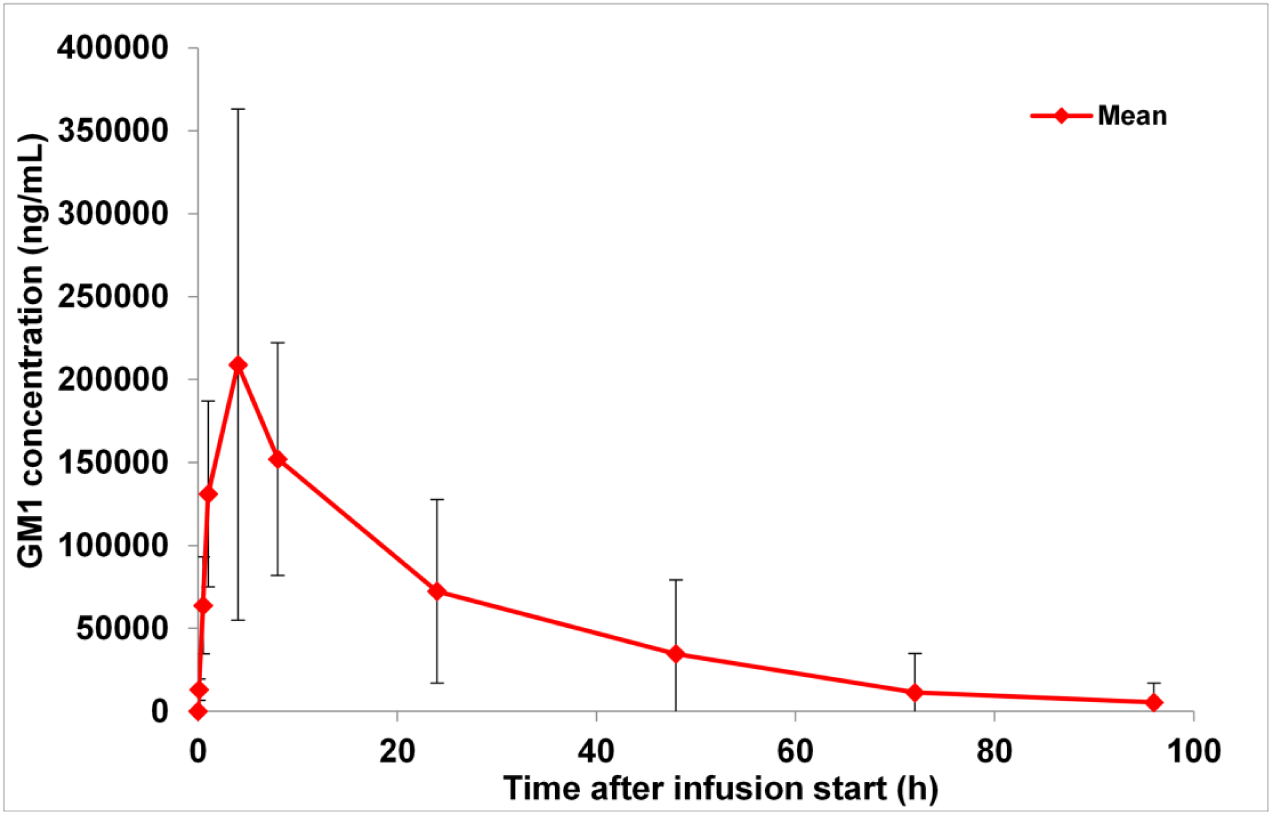
Mean (±SD) GM1 plasma concentration-time profiles after intravenous infusion administration of 720 mg GM1 (linear).

**Fig 3.**
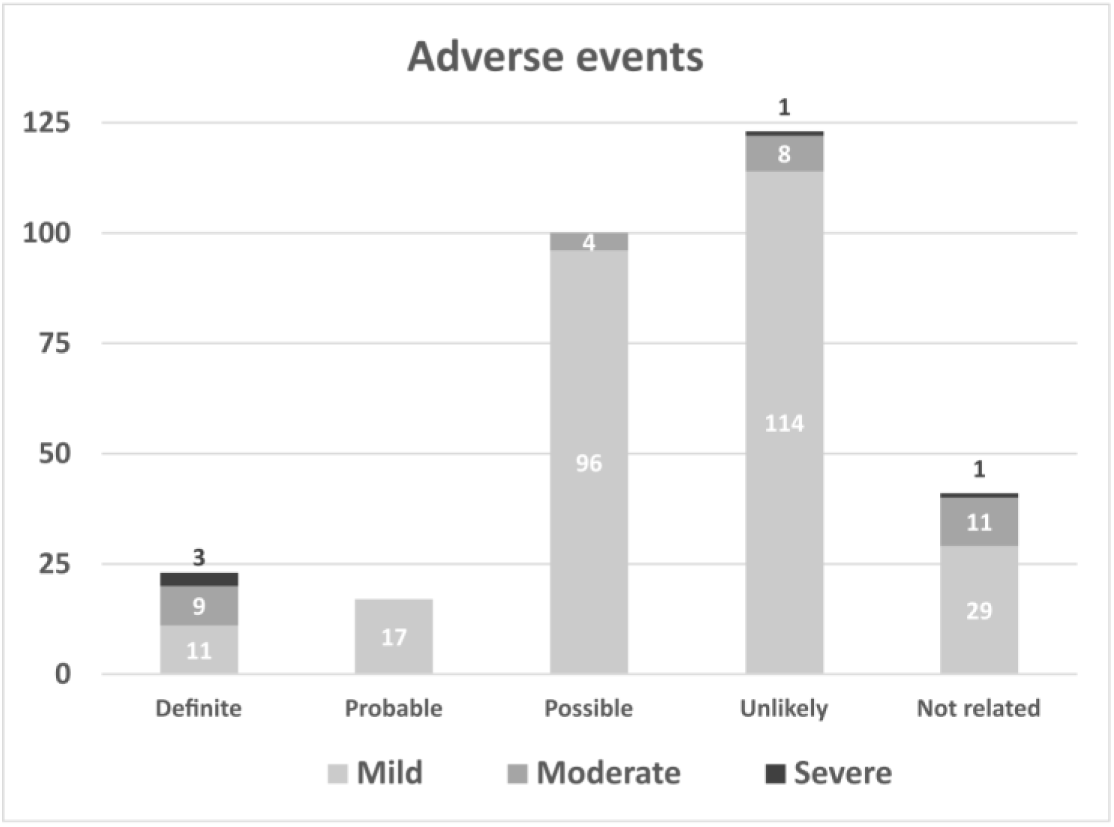
Observed adverse events (n = 304) during dose escalation and dose consolidation with Talineuren.

**Table 3.** The number of patients with acute infusion reaction and the presented symptoms early during the infusion.

| Acute infusion reaction | 2nd administration | 3rd administration | 4th administration |
| --- | --- | --- | --- |
| lower back pain | 7* | 2* | 1 |
| neck pain | 2* | 1* | 0 |
| itching, urticaria | 1 | 1 | 1 |
| nausea, chest pain | 2 | 0 | 0 |
| hypotension | 1 | 0 | 0 |
| thoracic congestion, plethora | 1 | 0 | 0 |
| eye flickering | 1 | 0 | 0 |

The number of patients presenting with acute symptoms early during the infusion is given.
\*one patient from the dose escalation group, all others are from the dose consolidation group.

Adverse events possibly related to TLN (Table 4) comprised 46 mild out of normal range measures of cholesterol, triglycerides, or apolipoprotein B. These observations were considered possibly related to TLN because the study drug contains cholesterol and sphingomyelin. However, lipid assessments were performed in a non-fasting state for which no ranges of normality were available. The lipid levels of all study participants were fluctuating over the study period but did not consistently change except for HDL cholesterol that was slightly but significantly reduced on all visits compared to baseline (see Appendix 2). In one patient, one observation of a slightly elevated level of anti-GM1 IgM antibodies was clinically asymptomatic and followed by normal levels in consecutive assessments. Many adverse events possibly related to TLN could also be explained as motor (e.g. muscle cramps, tremor, dysarthria, dysphagia) or non-motor (e.g., fatigue, sleep disorders, constipation, dysphoria) signs and symptoms of PD. During the four weeks pause after the dose escalation phase, one patient reported 25 adverse events of worsening parkinsonian motor and non-motor problems beyond baseline level. This rebound remitted spontaneously to baseline for all reported aspects before TLN was resumed in the dose consolidation phase. Mild tension headache, dizziness, nausea, and neck and lumbar pain were noted as possible infusion reaction symptoms.

**Table 4.** Adverse effects possibly related to the study treatment (n=100).

| Category of adverse events | n | System organ class terms | Preferred terms of grade 2 adverse events |
| --- | --- | --- | --- |
| Abnormal blood lipids | 46 | Investigations |  |
| Elevated anti-GM1-IgM antibodies | 1 | Investigations |  |
| Neurologic and psychiatric events | 25 | Nervous system disorders. Psychiatric disorders. Eye disorders. Renal and urinary disorders. Reproductive system and breast disorders |  |
| Gastrointestinal disorders | 7 | Gastrointestinal disorders | nausea |
| Asthenia and fatigue | 5 | General disorders and administration site conditions | fatigue |
| Possible infusion reactions | 7 | Injury, poisoning and procedural complications | procedural dizziness |
| Musculoskeletal problems | 8 | Musculoskeletal and connective tissue disorders | muscle spasms |
| Skin disorders | 1 | Skin and subcutaneous tissue disorders |  |

System organ class terms and preferred terms were used according to the MedDRA coding system for adverse effects.

**Table 5.** Adverse effects unlikely related to the study treatment (n=123).

| Category of adverse events | n | System organ class terms | Preferred terms of grade 2 and 3 adverse events |
| --- | --- | --- | --- |
| Out of range laboratory findings | 79 | Blood and lymphatic system disorders. Investigations.<br>Metabolism and nutrition disorders | increased blood potassium,<br>pseudohyperkalemia |
| Ear, nose and labyrinth disorder | 4 | Ear and labyrinth disorders.<br>Respiratory, thoracic and mediastinal disorders | acute vestibular syndrome |
| Gastrointestinal disorders | 18 | Gastrointestinal disorders | nausea, upper abdominal pain |
| Infections and infestations | 2 | Infections and infestations | post viral fatigue syndrome |
| Cardiovascular disorders | 2 | Cardiac disorders. Eye disorders | amaurosis fugax |
| Musculoskeletal and connective tissue disorders | 9 | Musculoskeletal and connective tissue disorders | back pain |
| Nervous system and psychiatric disorders | 8 | Nervous system disorders. Psychiatric disorders | tension headache |
| Skin and subcutaneous tissue disorders | 1 | Skin and subcutaneous tissue disorders |  |

System organ class terms and preferred terms were used according to the MedDRA coding system for adverse effects.

Adverse events unlikely related to TLN (table 4) comprised mainly lab results mildly outside of the normal range. Two measures of hyperkalaemia (5.5 and 5.8 mmol/l) were clinically asymptomatic and occurred without other lab abnormalities; an artefact due to erroneous sample handling was assumed. A self-remitting episode of acute vestibular vertigo was the only severe adverse event considered unlikely to be related to TLN.

### Secondary outcome measures

At an early stage, 1 patient discontinued the study and was removed from the efficacy analysis, meaning that the present analysis was performed on 11 patients. At the final assessment, parkinsonian motor signs off medication improved 6.73±7.16 points (Pval = 0.01 ) on the MDS-UPDRS 3 assessed after 12 hours of withdrawal of dopaminergic medication (see Table 6). This corresponded with the observation that pausing medication for the final assessment was much better tolerated than at baseline. With medication (on state), no significant change in motor signs was found during the study. TLN treatment did not result in a consistent and clinically relevant immediate beneficial effect on parkinsonian motor signs (data not shown). Motor complications of dopaminergic treatment (MDS-UPDRS 4) did not change significantly during the study. There were fewer non-motor symptoms at follow-up compared to baseline as assessed with the NMS-Quest (8.64 vs. 4.63, Pval = 0.009). Non-motor parkinsonian symptoms (MDS-UPDRS 1) and activities of daily living in the best condition (MDS-UPDRS 2 best) were significantly improved at follow-up compared to baseline (mean difference from baseline -1.91, pval = 0.057 and -2.82, pval = 0.02; respectively).

**Table 6.**
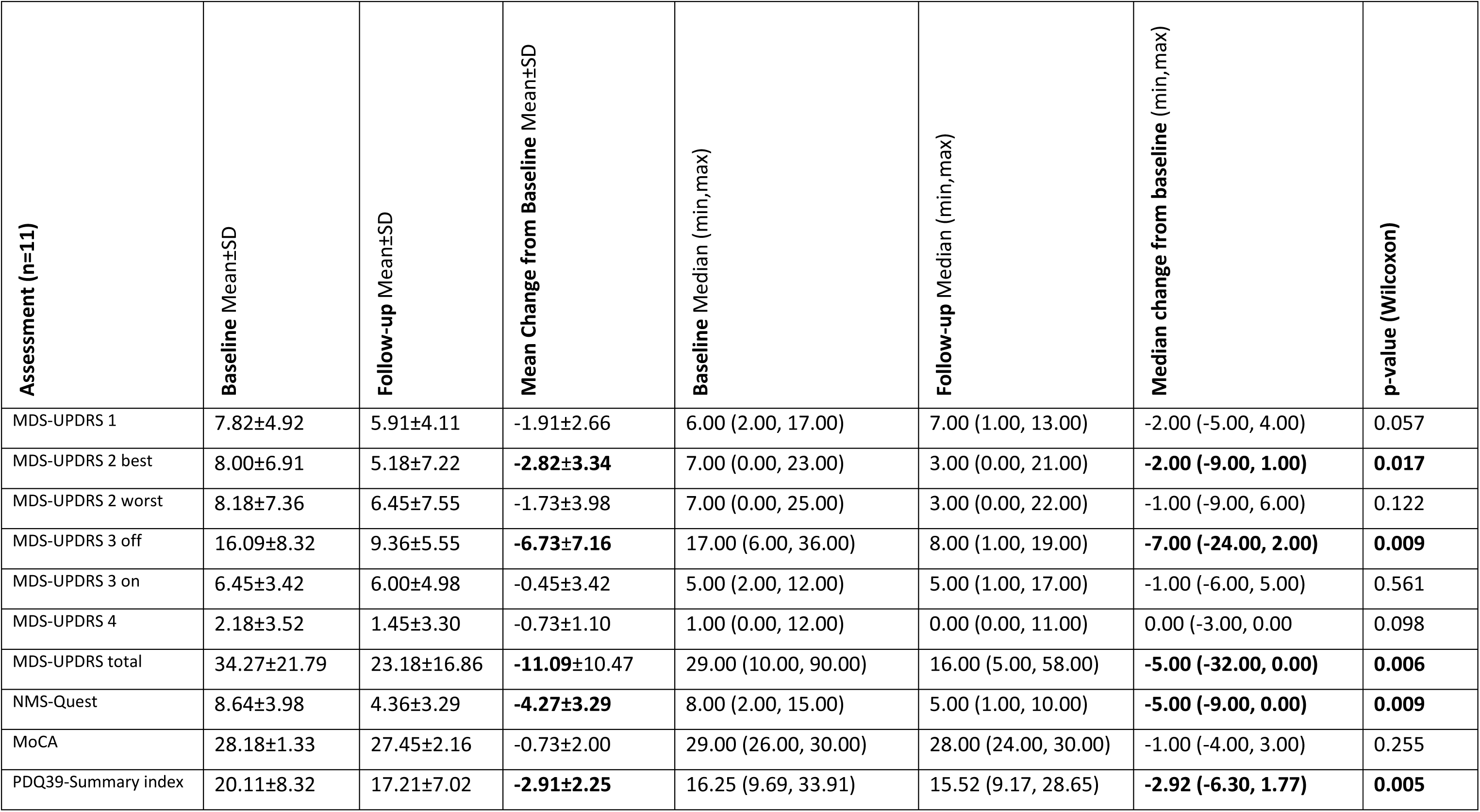
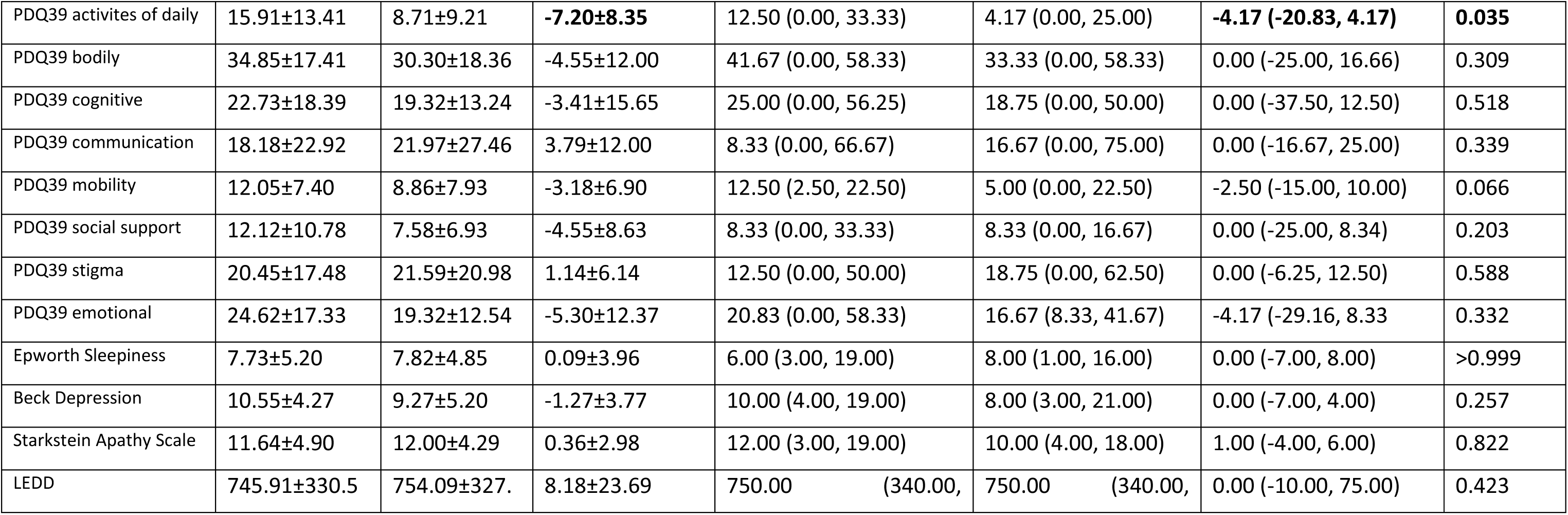
Motor signs and non-motor symptoms, quality of life, and LEDD for the 11 patients.

| Assessment (n=11) | Baseline Mean±SD | Follow-up Mean±SD | Mean Change from Baseline Mean±SD | Baseline Median (min,max) | Follow-up Median (min,max) | Median change from baseline (min,max) | p-value (Wilcoxon) |
| --- | --- | --- | --- | --- | --- | --- | --- |
| MDS-UPDRS 1 | 7.82±4.92 | 5.91±4.11 | -1.91±2.66 | 6.00 (2.00, 17.00) | 7.00 (1.00, 13.00) | -2.00 (-5.00, 4.00) | 0.057 |
| MDS-UPDRS 2 best | 8.00±6.91 | 5.18±7.22 | **-2.82±3.34** | 7.00 (0.00, 23.00) | 3.00 (0.00, 21.00) | **-2.00 (-9.00, 1.00)** | **0.017** |
| MDS-UPDRS 2 worst | 8.18±7.36 | 6.45±7.55 | -1.73±3.98 | 7.00 (0.00, 25.00) | 3.00 (0.00, 22.00) | -1.00 (-9.00, 6.00) | 0.122 |
| MDS-UPDRS 3 off | 16.09±8.32 | 9.36±5.55 | **-6.73±7.16** | 17.00 (6.00, 36.00) | 8.00 (1.00, 19.00) | **-7.00 (-24.00, 2.00)** | **0.009** |
| MDS-UPDRS 3 on | 6.45±3.42 | 6.00±4.98 | -0.45±3.42 | 5.00 (2.00, 12.00) | 5.00 (1.00, 17.00) | -1.00 (-6.00, 5.00) | 0.561 |
| MDS-UPDRS 4 | 2.18±3.52 | 1.45±3.30 | -0.73±1.10 | 1.00 (0.00, 12.00) | 0.00 (0.00, 11.00) | 0.00 (-3.00, 0.00) | 0.098 |
| MDS-UPDRS total | 34.27±21.79 | 23.18±16.86 | **-11.09±10.47** | 29.00 (10.00, 90.00) | 16.00 (5.00, 58.00) | **-5.00 (-32.00, 0.00)** | **0.006** |
| NMS-Quest | 8.64±3.98 | 4.36±3.29 | **-4.27±3.29** | 8.00 (2.00, 15.00) | 5.00 (1.00, 10.00) | **-5.00 (-9.00, 0.00)** | **0.009** |
| MoCA | 28.18±1.33 | 27.45±2.16 | -0.73±2.00 | 29.00 (26.00, 30.00) | 28.00 (24.00, 30.00) | -1.00 (-4.00, 3.00) | 0.255 |
| PDQ39-Summary index | 20.11±8.32 | 17.21±7.02 | **-2.91±2.25** | 16.25 (9.69, 33.91) | 15.52 (9.17, 28.65) | **-2.92 (-6.30, 1.77)** | **0.005** |
| PDQ39 activities of daily | 15.91±13.41 | 8.71±9.21 | **-7.20±8.35** | 12.50 (0.00, 33.33) | 4.17 (0.00, 25.00) | **-4.17 (-20.83, 4.17)** | **0.035** |
| PDQ39 bodily | 34.85±17.41 | 30.30±18.36 | -4.55±12.00 | 41.67 (0.00, 58.33) | 33.33 (0.00, 58.33) | 0.00 (-25.00, 16.66) | 0.309 |
| PDQ39 cognitive | 22.73±18.39 | 19.32±13.24 | -3.41±15.65 | 25.00 (0.00, 56.25) | 18.75 (0.00, 50.00) | 0.00 (-37.50, 12.50) | 0.518 |
| PDQ39 communication | 18.18±22.92 | 21.97±27.46 | 3.79±12.00 | 8.33 (0.00, 66.67) | 16.67 (0.00, 75.00) | 0.00 (-16.67, 25.00) | 0.339 |
| PDQ39 mobility | 12.05±7.40 | 8.86±7.93 | -3.18±6.90 | 12.50 (2.50, 22.50) | 5.00 (0.00, 22.50) | -2.50 (-15.00, 10.00) | 0.066 |
| PDQ39 social support | 12.12±10.78 | 7.58±6.93 | -4.55±8.63 | 8.33 (0.00, 33.33) | 8.33 (0.00, 16.67) | 0.00 (-25.00, 8.34) | 0.203 |
| PDQ39 stigma | 20.45±17.48 | 21.59±20.98 | 1.14±6.14 | 12.50 (0.00, 50.00) | 18.75 (0.00, 62.50) | 0.00 (-6.25, 12.50) | 0.588 |
| PDQ39 emotional | 24.62±17.33 | 19.32±12.54 | -5.30±12.37 | 20.83 (0.00, 58.33) | 16.67 (8.33, 41.67) | -4.17 (-29.16, 8.33) | 0.332 |
| Epworth Sleepiness | 7.73±5.20 | 7.82±4.85 | 0.09±3.96 | 6.00 (3.00, 19.00) | 8.00 (1.00, 16.00) | 0.00 (-7.00, 8.00) | >0.999 |
| Beck Depression | 10.55±4.27 | 9.27±5.20 | -1.27±3.77 | 10.00 (4.00, 19.00) | 8.00 (3.00, 21.00) | 0.00 (-7.00, 4.00) | 0.257 |
| Starkstein Apathy Scale | 11.64±4.90 | 12.00±4.29 | 0.36±2.98 | 12.00 (3.00, 19.00) | 10.00 (4.00, 18.00) | 1.00 (-4.00, 6.00) | 0.822 |
| LEDD | 745.91±330.5 | 754.09±327. | 8.18±23.69 | 750.00 (340.00, | 750.00 (340.00, | 0.00 (-10.00, 75.00) | 0.423 |

The total MDS-UPDRS (including the MDS-UPDRS 2 in the worst condition and the MDS-UPDRS 3 off medication) improved by 11.1±10.47 from baseline to follow-up (pval = 0.006).

Quality of life related to PD as measured with the PDQ39-SI improved by 2.91±2.25 (pval = 0.005), mainly driven by an improvement of quality of life for activities of daily living. The other subdomains of the PDQ39 did not improve significantly. No significant change in mood (BDI), motivation (Starkstein Apathy Scale), overall cognition (MoCA), and dopaminergic medication (levodopa equivalent daily dose [LEDD]) was observed.

During dose-escalation, one patient reported a general effect of well-being and more energy that lasted longer with increasing doses up to 1 week with GM1 720 mg. All patients reported mild to moderate reduction of the previously perceived beneficial effects during the month between the last infusion and the follow-up safety assessment. No lasting worsening beyond baseline was observed after one month’s withdrawal of TLN. In the first three patients, total GM1 dose was resumed at 720 mg weekly after a month’s pause following dose escalation; within one or two infusions the previous level of well-being was re-attained. All patients emphatically requested to prolong TLN treatment at the end of the study. An amendment for study prolongation was therefore submitted.

## Discussion

In this open phase I clinical safety trial 12 PD patients received weekly infusions with liposomal GM1 for 8 to 22 weeks. Tolerability was overall very good with no or mild unspecific adverse reactions to the infusion. Some fatigue and dizziness occurred repeatedly in some but not all patients after the infusion and lasted for a few hours, so a causal link to TLN seems probable. However, there were acute infusion reactions in 7 of the 12 patients at the 2nd, 3rd, or 4th exposure to TLN. The intensity was mild to moderate and in one case severe leading to exclusion from further study treatment. These reactions were most pronounced at second exposure, consisted mostly of neck and lumbar back pain, and swiftly abated within minutes after TLN infusions were paused. An allergic IgE-mediated anaphylactic reaction seems highly unlikely as re-exposure did not lead to reappearance of the symptoms but to habituation. Moreover, tryptase was measured before and after an acute infusion reaction with urticaria at the 3rd exposure in one patient and remained stable in the normal range. We therefore considered a complement activation related pseudo-allergy (CARPA) to the liposome as an explanation for the acute infusion reactions (23). However, CARPAs typically occur with the first exposure and habituate thereafter, whereas in our patients the first exposure was unproblematic in all 12 patients. Moreover, the clinical manifestation of CARPAs is not typically reported as back pain which was the main manifestation in our patients. The occurrence of the acute infusion reaction only at the 2nd administration is suggestive of a sensitization to TLN, however, the unproblematic tolerance of re-exposure makes autoantibodies to GM1 an unlikely explanation, especially as anti-GM1-IgM levels remained normal throughout the study in these patients and did not increase. Although we cannot explain the mechanism of the observed acute infusion reactions, slowing the infusion rate and possibly progressively increasing doses over the first weeks of treatment with TLN seem to prevent such reactions. Free GM1 has been administered intravenously for over three decades in several neurological disorders for up to 2500 mg/d without observing acute infusion reactions (24). In particular in the randomized placebo-controlled trial with subcutaneous free GM1 patients received a single intravenous loading dose of 1000 mg GM1. The subsequent subcutaneous administration of GM1 in this study was a route of administration with a much higher risk of triggering an allergic reaction to GM1 than intravenous administration, but no allergic side effects were reported. Therefore, we suspect the acute infusion reactions in our patients to be related to the liposomal carrier of GM1. Further analyses of possible mechanisms for an acute reaction to TLN are ongoing outside of this trial. Other safety assessments of TLN will include lab analyses of blood samples drawn after potential further acute infusion reactions to assess tryptase, thromboxanes, complement factors, activation of factor XII, and measures of basophile activation. However, we try to completely avoid further adverse reactions of this nature by gradually increasing the dose of TLN over the first weeks and by starting the speed of infusion at a very low rate and ramping up as tolerated, especially for the 2nd and 3rd administration of TLN.

Although hyperlipidemia was present in some of our patients already at baseline, there was no consistent increase of lipid levels throughout the trial as had been observed with the administration of very high doses of free GM1 previously (24). Hyperlipidemia was likely overestimated in our patients as blood was drawn in non-fasting conditions. However, HDL cholesterol was slightly but significantly lowered under TLN treatment which may consist in a vascular risk factor. There were no adverse events related to atherosclerosis in this study, but the observation period was very short. For more conclusive results, lipid levels must be assessed systematically and under standardized fasting conditions in patients treated with TLN in future studies.

Although this is an uncontrolled phase I safety trial, we gathered exploratory data on the possible therapeutic effects of TLN as a basis for power calculations for a therapeutic trial. In terms of PD symptoms, all patients felt stable or better over the study period. Improvements of sleep quality, motivation, sense of smell, and overall energy were reported, gradually occurring over several weeks. The drug withdrawal for the second levodopa challenge test was much better tolerated by most patients than the first, and several participants reported that accidental omission of a dose of levodopa was much less perceived than before the treatment with TLN. These encouraging observations are reflected in an improvement of non-motor (MDS-UPDRS-1, NMS-Quest) and motor aspects of experiences of daily living (MDS-UPDRS-2), and of parkinsonian motor signs (MDS-UPDRS- 3) resulting in an improvement of disease-related quality of life (PDQ-39). The observed improvements of the MDS-UPDRS-1 and MDS-UPDRS-2 of 1.91 and 2.82 points are below the minimal clinically relevant difference of 2.64 and 3.05 for these scales (25). However, for the motor signs, the minimal clinical improvement on the MDS-UPDRS-3 has been suggested at 3.5 points (26), and our patients improved by 6.73 points. The minimal clinically relevant improvement on the MDS-UPDRS total score has been estimated at 7.1 points (27), whereas our patients improved by 11.09 points. However, for disease-related quality of life measured by the PDQ-39 the minimal clinically important improvement of 4.72% (28) is not reached with 2.9% observed in our patients. These exploratory observations are encouraging, especially as they uniformly point towards an improvement on different established scales despite a very small number of patients. The observed improvements occurred gradually over the study period. No immediate effect on motor signs was found in the ratings before and after TLN infusions. A rapid symptomatic beneficial effect of TLN on parkinsonian signs and symptoms is therefore most unlikely. Moreover, as there was no selection of patients based on minimum disease severity, regression to the mean is unlikely to explain the observed improvement in this study. If there is indeed a therapeutic effect of TLN, either a long-term symptomatic effect or a disease-modifying effect seems possible. As the annual progression of PD in its natural course has been estimated at 5.05 points in the untreated and 2.13 points in the treated condition for the sum of the MDS-UPDRS-2 and -3 (29), the observed improvement among our patients may point to a long-term symptomatic or even a neurorestorative effect. Indeed, GM1 can stabilize alpha-synuclein (7) and has a neurorestorative effect on neurons in animal models of PD (30). Suffering neurons in patients with PD may therefore be rescued by GM1 treatment, and a liposomal formulation may be an effective approach to deliver GM1 into the central and peripheral nervous system. This may explain a gradual and long-lasting reparative effect of TLN despite its short plasma half-life of 12.6h. However, all this remains speculative at present. As this trial is not designed to show a therapeutic effect of TLN given the open design without placebo control, a placebo effect may explain the observed improvements. An adequately powered randomized placebo-controlled trial of TLN in PD is needed to further explore this promising new treatment.

## Data Availability

Combinations of the demographic data could possibly identify participants and thereby compromise participant privacy. Thus, per-patient details on demographics and eligibility criteria cannot be shared. All remaining relevant data are within the manuscript and its Supporting Information files.

## Acknowledgments

MedDRA® trademark is registered by ICH.

## Trial Registration

The NEON trial is registered at the US National Institutes of Health (ClinicalTrials.gov) #NCT04976127 and in the Swiss National Clinical Trials Portal (SNCTP000004631) **Funding:** NEON was fully funded by InnoMedica Switzerland AG.

## Supporting information

**<u>S1 Checklist.</u> Transparent Reporting of Evaluations with Nonrandomized Designs (TREND)**

**statement checklist.**

**S2 Text. Clinical trial protocol.**

## Appendix 1 Laboratory analysis list

Sodium, potassium, calcium, phosphate, creatinine, urea, albumin, total protein, cystatin C, lactate dehydrogenase, ferritin, HbA1c, ALT, AST, total bilirubin, pancreatic amylase, alkaline phosphatase, cholesterol (total, LDL, HDL), apolipoprotein B, differentiated red and white blood cell count, thrombocytes, ganglioside GM1 IgG antibodies

## Appendix 2: Observed lab values and weekly change

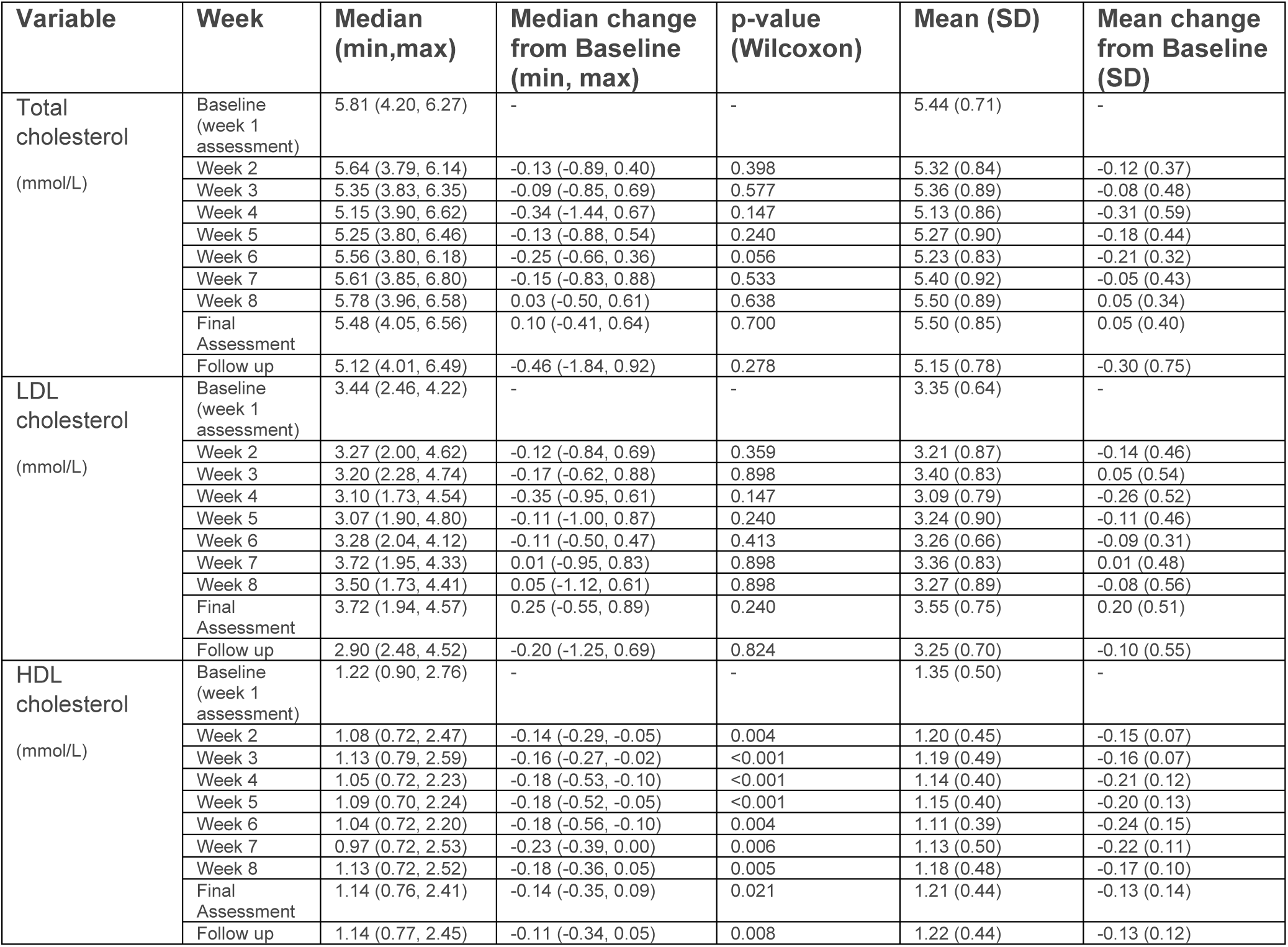
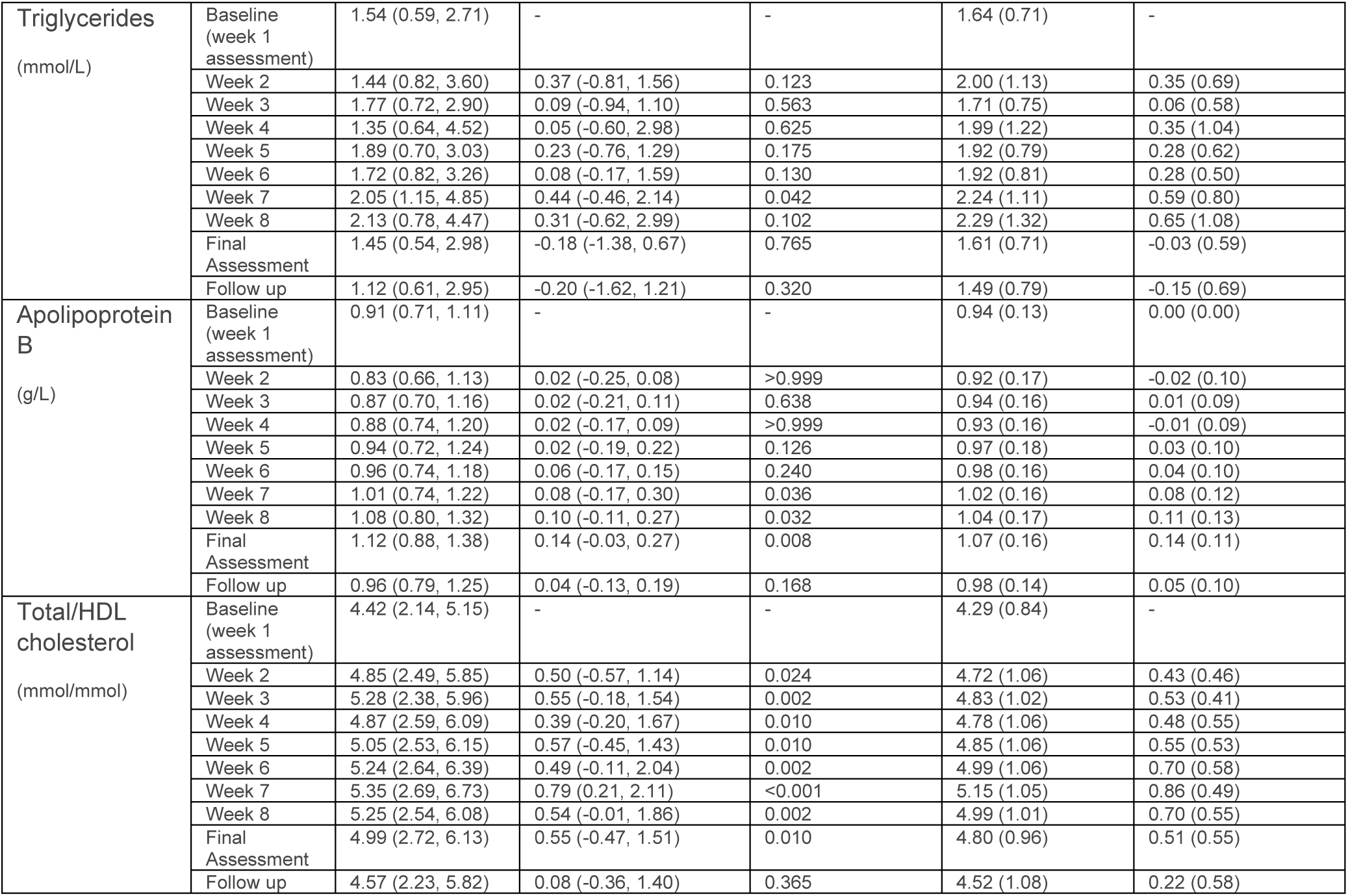

## Notes

### Competing Interest Statement

SH, SL, PP, R-PC, CP, PCB and CK are or have been employees at InnoMedica Schweiz AG and may additionally hold shares. SH and CP are board members of InnoMedica Schweiz AG. MS has received honoraria as a consultant for Medtronic.

### Clinical Trial

Trial number: NCT04976127 https://clinicaltrials.gov/study/NCT04976127

### Funding Statement

Yes

### Author Declarations

Ethics committee: Kantonale Ethikkommission Bern

## References

1. Postuma RB, Berg D, Stern M, Poewe W, Olanow CW, Oertel W, et al. MDS clinical diagnostic criteria for Parkinson’s disease. Mov Disord [Internet]. 2015 Oct 1 [cited 2023 Jun 20];30(12):1591–601. Available from: https://pubmed.ncbi.nlm.nih.gov/26474316/

2. Berg D, Borghammer P, Fereshtehnejad SM, Heinzel S, Horsager J, Schaeffer E, et al. Prodromal Parkinson disease subtypes - key to understanding heterogeneity. Nat Rev Neurol [Internet]. 2021 Jun 1 [cited 2023 Jun 20];17(6):349–61. Available from: https://pubmed.ncbi.nlm.nih.gov/33879872/

3. Braak H, Del Tredici K, Rüb U, De Vos RAI, Jansen Steur ENH, Braak E. Staging of brain pathology related to sporadic Parkinson’s disease. Neurobiol Aging [Internet]. 2003 Mar [cited 2023 Jun 20];24(2):197–211. Available from: https://pubmed.ncbi.nlm.nih.gov/12498954/

4. Rietdijk CD, Perez-Pardo P, Garssen J, van Wezel RJA, Kraneveld AD. Exploring Braak’s Hypothesis of Parkinson’s Disease. Front Neurol [Internet]. 2017 Feb 13 [cited 2023 Jun 20];8(FEB). Available from: https://pubmed.ncbi.nlm.nih.gov/28243222/

5. McFarthing K, Rafaloff G, Baptista M, Mursaleen L, Fuest R, Wyse RK, et al. Parkinson’s Disease Drug Therapies in the Clinical Trial Pipeline: 2022 Update. J Parkinsons Dis [Internet]. 2022 [cited 2023 Jun 20];12(4):1073–82. Available from: https://pubmed.ncbi.nlm.nih.gov/35527571/

6. te Vruchte D, Sturchio A, Priestman DA, Tsitsi P, Hertz E, Andréasson M, et al. Glycosphingolipid Changes in Plasma in Parkinson’s Disease Independent of Glucosylceramide Levels. Mov Disord [Internet]. 2022 Oct 1 [cited 2023 Jun 20];37(10):2129–34. Available from: https://pubmed.ncbi.nlm.nih.gov/35876461/

7. Sonnino S. The relationship between depletion of brain GM1 ganglioside and Parkinson’s disease. FEBS Open Bio [Internet]. 2023 [cited 2023 Jun 20]; Available from: https://pubmed.ncbi.nlm.nih.gov/36638010/

8. Chowdhury S, Ledeen R. The Key Role of GM1 Ganglioside in Parkinson’s Disease. Biomolecules [Internet]. 2022 Jan 21 [cited 2023 Jun 20];12(2). Available from: https://pubmed.ncbi.nlm.nih.gov/35204675/

9. Schneider JS, Pope A, Simpson K, Taggart J, Smith MG, DiStefano L. Recovery from experimental parkinsonism in primates with GM1 ganglioside treatment. Science [Internet]. 1992 [cited 2023 Jun 20];256(5058):843–6. Available from: https://pubmed.ncbi.nlm.nih.gov/1350379/

10. Schneider JS, Gollomp SM, Sendek S, Colcher A, Cambi F, Du W, et al. A Randomized, Controlled, Delayed Start Trial of GM1 Ganglioside in Treated Parkinson’s Disease Patients Jay. J Neurol Sci [Internet]. 2013;324(1–2):140–8. Available from: 10.1016/j.jns.2012.10.024

11. Schneider JS, Gollomp SM, Sendek S, Colcher A, Cambi F, Du W. A randomized, controlled, delayed start trial of GM1 ganglioside in treated Parkinson’s disease patients. J Neurol Sci [Internet]. 2013 Jan 15 [cited 2023 Jun 20];324(1–2):140–8. Available from: https://pubmed.ncbi.nlm.nih.gov/23199590/

12. Hughes AJ, Daniel SE, Kilford L, Lees AJ. Accuracy of clinical diagnosis of idiopathic Parkinson’s disease: a clinico-pathological study of 100 cases. J Neurol Neurosurg Psychiatry [Internet]. 1992 [cited 2023 Jun 20];55(3):181–4. Available from: https://pubmed.ncbi.nlm.nih.gov/1564476/

13. Hoehn MM, Yahr MD. Parkinsonism: onset, progression and mortality. Neurology [Internet]. 1967 [cited 2023 Jun 20];17(5):427–42. Available from: https://pubmed.ncbi.nlm.nih.gov/6067254/

14. Nasreddine ZS, Phillips NA, Bédirian V, Charbonneau S, Whitehead V, Collin I, et al. The Montreal Cognitive Assessment, MoCA: a brief screening tool for mild cognitive impairment. J Am Geriatr Soc [Internet]. 2005 [cited 2023 Jun 20];53(4):695–9. Available from: https://pubmed.ncbi.nlm.nih.gov/15817019/

15. Goetz CG, Tilley BC, Shaftman SR, Stebbins GT, Fahn S, Martinez-Martin P, et al. Movement Disorder Society-sponsored revision of the Unified Parkinson’s Disease Rating Scale (MDS-UPDRS): scale presentation and clinimetric testing results. Mov Disord [Internet]. 2008 Nov 15 [cited 2023 Jun 20];23(15):2129–70. Available from: https://pubmed.ncbi.nlm.nih.gov/19025984/

16. Nyholm D, Jost WH. An updated calculator for determining levodopa-equivalent dose. Neurol Res Pract [Internet]. 2021 Dec 1 [cited 2023 Jun 20];3(1). Available from: https://pubmed.ncbi.nlm.nih.gov/34689840/

17. Chaudhuri KR, Martinez-Martin P, Schapira AHV, Stocchi F, Sethi K, Odin P, et al. International multicenter pilot study of the first comprehensive self-completed nonmotor symptoms questionnaire for Parkinson’s disease: the NMSQuest study. Mov Disord [Internet]. 2006 [cited 2023 Jun 20];21(7):916–23. Available from: https://pubmed.ncbi.nlm.nih.gov/16547944/

18. Peto V, Jenkinson C, Fitzpatrick R, Greenhall R. The development and validation of a short measure of functioning and well being for individuals with Parkinson’s disease. Qual Life Res [Internet]. 1995 Jun [cited 2023 Jun 20];4(3):241–8. Available from: https://pubmed.ncbi.nlm.nih.gov/7613534/

19. Johns MW. A new method for measuring daytime sleepiness: the Epworth sleepiness scale. Sleep [Internet]. 1991 [cited 2023 Jun 20];14(6):540–5. Available from: https://pubmed.ncbi.nlm.nih.gov/1798888/

20. Nasreddine ZS, Phillips NA, Bédirian V, Charbonneau S, Whitehead V, Collin I, et al. The Montreal Cognitive Assessment, MoCA: a brief screening tool for mild cognitive impairment. J Am Geriatr Soc [Internet]. 2005 [cited 2023 Jun 20];53(4):695–9. Available from: https://pubmed.ncbi.nlm.nih.gov/15817019/

21. Beck AT, Ward CH, Mendelson M, Mock J, Erbaugh J. An inventory for measuring depression. Arch Gen Psychiatry [Internet]. 1961 [cited 2023 Jun 20];4(6):561–71. Available from: https://pubmed.ncbi.nlm.nih.gov/13688369/

22. Starkstein SE, Mayberg HS, Preziosi TJ, Andrezejewski P, Leiguarda R, Robinson RG. Reliability, validity, and clinical correlates of apathy in Parkinson’s disease. J Neuropsychiatry Clin Neurosci [Internet]. 1992 [cited 2023 Jun 20];4(2):134–9. Available from: https://pubmed.ncbi.nlm.nih.gov/1627973/

23. Szebeni J, Simberg D, González-Fernández Á, Barenholz Y, Dobrovolskaia MA. Roadmap and strategy for overcoming infusion reactions to nanomedicines. Nat Nanotechnol [Internet]. 2018 Dec 1 [cited 2023 Jun 20];13(12):1100–8. Available from: https://pubmed.ncbi.nlm.nih.gov/30348955/

24. Roberts JW, Hoeg JM, Maral Mouradian M, Linfante I, Chase TN. Iatrogenic hyperlipidaemia with GM1 ganglioside. Lancet [Internet]. 1993 Jul 10 [cited 2023 Jun 20];342(8863):115. Available from: https://pubmed.ncbi.nlm.nih.gov/8100881/

25. Horváth K, Aschermann Z, Kovács M, Makkos A, Harmat M, Janszky J, et al. Minimal clinically important differences for the experiences of daily living parts of movement disorder society-sponsored unified Parkinson’s disease rating scale. Mov Disord [Internet]. 2017 May 1 [cited 2023 Jun 20];32(5):789–93. Available from: https://pubmed.ncbi.nlm.nih.gov/28218413/

26. Horváth K, Aschermann Z, Ács P, Deli G, Janszky J, Komoly S, et al. Minimal clinically important difference on the Motor Examination part of MDS-UPDRS. Parkinsonism Relat Disord [Internet]. 2015 Dec 1 [cited 2023 Jun 20];21(12):1421–6. Available from: https://pubmed.ncbi.nlm.nih.gov/26578041/

27. Makkos A, Kovács M, Aschermann Z, Harmat M, Janszky J, Karádi K, et al. Are the MDS-UPDRS-Based Composite Scores Clinically Applicable? Mov Disord [Internet]. 2018 May 1 [cited 2023 Jun 20];33(5):835–9. Available from: https://pubmed.ncbi.nlm.nih.gov/29488318/

28. Horváth K, Aschermann Z, Kovács M, Makkos A, Harmat M, Janszky J, et al. Changes in Quality of Life in Parkinson’s Disease: How Large Must They Be to Be Relevant? Neuroepidemiology [Internet]. 2017 Jun 1 [cited 2023 Jun 20];48(1–2):1–8. Available from: https://pubmed.ncbi.nlm.nih.gov/28161701/

29. Latourelle JC, Beste MT, Hadzi TC, Miller RE, Oppenheim JN, Valko MP, et al. Large-scale identification of clinical and genetic predictors of motor progression in patients with newly diagnosed Parkinson’s disease: a longitudinal cohort study and validation. Lancet Neurol [Internet]. 2017 Nov 1 [cited 2023 Jun 20];16(11):908–16. Available from: https://pubmed.ncbi.nlm.nih.gov/28958801/

30. Schneider JS, Aras R, Williams CK, Koprich JB, Brotchie JM, Singh V. GM1 Ganglioside Modifies α-Synuclein Toxicity and is Neuroprotective in a Rat α-Synuclein Model of Parkinson’s Disease. Sci Rep [Internet]. 2019 Dec 1 [cited 2023 Jun 20];9(1). Available from: https://pubmed.ncbi.nlm.nih.gov/31182727/

